# A computational genetic- and transcriptomics-based study nominates drug repurposing candidates for the treatment of chronic pain

**DOI:** 10.1101/2025.03.07.25323591

**Authors:** Alanna C. Cote, Keira J.A. Johnston, Carina Seah, Hannah Young, Laura M. Huckins

## Abstract

Chronic pain, defined as pain that persists for greater than three months, is a common, understudied condition that affect an estimated 20-30% of the population^1,2^. Despite a high prevalence and distressing physical and psychological symptoms, research is lacking in appropriate long-term pharmaceutical treatment for chronic pain, and chronic pain persists at high rates even with intervention. Recent genome-wide association studies (GWAS) of chronic pain indicate that chronic pain can be studied as a distinct neuropsychiatric illness with genetic risk^3,4^. Here we develop a genetics-informed framework to identify new drug repurposing candidates for chronic pain. We first use a functional genomics approach to drug repurposing, called signature mapping, to identify drug repurposing candidates for chronic pain^5^. In a signature mapping analysis, the transcriptomic effects of disease and drug perturbations are compared, and drugs with opposite effects on gene expression as the disease are nominated as therapeutic candidates. Then we further investigate therapeutic avenues through causal inference using two-sample Mendelian randomization analysis, leveraging GWAS of chronic pain and GWAS of medication use across 23 drug classes. This approach, driven by information from genome- and transcriptome-wide associations with chronic pain, can yield additional insights beyond the single receptor- or molecule-level drug discovery investigation in traditional drug development pipelines.

## Introduction

Chronic pain, broadly defined as pain lasting longer than three months, is a common and understudied condition that affects an estimated 20-30% of the population^1,2^. Chronic pain constitutes a considerable disease burden; chronic pain conditions such as low back or neck pain, musculoskeletal disorders, and migraine are among the top ten leading causes of disability and injury^6^, and recent incidence estimates of chronic pain exceed that of other well-known conditions like diabetes, depression, and hypertension^7^.

Chronic pain is a highly complex condition with dynamic biological, social, and psychological impacts on disease risk and long-term outcomes; characteristics like depression, anxiety, poor sleep, or poor social support systems can act as both a risk factor for and consequence of chronic pain development^8,9^. Although many specific diseases can cause chronic pain, there are multiple lines of evidence suggesting that the study of chronic pain as a primary complex disease trait is also warranted^8^. Many chronic pain conditions co-occur, with pain commonly reported in multiple areas of the body and considerable comorbidity observed between seemingly distinct pain conditions, suggesting some underlying shared mechanisms among chronic pain disorders^10^. In addition, the relationship between the degree of acute injury or pain and later development of chronic pain is not well defined. Only a subset of patients with a particular disease go on to develop chronic pain, and for some conditions such as osteoarthritis there is no discernable relationship between pain and measurable tissue damage^11–13^. In the last ten years the first genome-wide association studies (GWAS) of chronic pain indicate that chronic pain is a complex trait with distinct genetic risk, with moderate SNP-heritability of about 10-20%^3,4^.

Chronic pain is most effectively treated using an interdisciplinary approach, with a combination of medication and non-drug alternatives such as cognitive behavioral therapy, exercise, or physical therapy^9^. Unfortunately, these nonpharmacological treatments are not always covered by insurance, and thus not easily accessible to individuals of lower socioeconomic status seeking chronic pain treatment. In terms of medication, chronic pain has been historically treated with opioids, but given the risk of addiction with opioid treatment, recent CDC and FDA guidelines emphasize non-opioid alternatives, including gabapentin, pregabalin, non-steroidal anti-inflammatory drugs (NSAIDs), antidepressants, anticonvulsants, and topical treatments such as lidocaine and capsaicin. There is insufficient evidence supporting the sustained effectiveness of these drugs for long-term treatment of chronic pain and these medications show small to moderate efficacy in short-term trials^10,14,15^, highlighting a continued need for drug alternatives.

In this study we use a validated functional genomics approach to drug repurposing, called signature mapping, to identify novel drug repurposing candidates for chronic pain, and investigate causal effects of broader drug classes on chronic pain in a complementary Mendelian randomization analysis. In a signature mapping analysis the transcriptomic effects of disease and drug perturbations are compared, and drugs with opposite effects on gene expression as the disease are nominated as therapeutic candidates^5^. There are few well-powered human differential expression studies of chronic pain or pain-related conditions (∼n < 150 cases)^16–20^. Instead, we perform summary-based transcriptome-wide association studies of chronic pain, measuring the association of estimated gene expression with the presence and burden of chronic multisite pain^3,4^. Using these disease signatures and a published differential expression study of acute pain resolution^21^, we perform a signature mapping analysis measuring against a public resource of drug effects on expression across cell lines^22^. We apply a novel ensemble score, combining both enrichment- and similarity-based signature mapping scores, to quantify the difference between drug and disease effects on the transcriptome. Then we further investigate therapeutic avenues through causal inference using two-sample Mendelian randomization analysis, leveraging GWAS of chronic pain and GWAS of medication use across 23 drug classes.

There are multiple motivations for applying this genome- and transcriptome-wide framework in the context of chronic pain. Signature mapping-based drug repurposing can be a fast and cost-effective alternative to drug discovery for the pharmaceutical treatment of complex diseases, and despite its simple design has shown success in prior diseases with varying levels of statistical and experimental follow-up^5,23–27^. Drugs with genetic evidence are more likely to proceed to late-stage clinical trials^28,29^. Pain is a complex phenotype that is not likely to be sufficiently treated using the single gene or molecular target approaches common in traditional drug discovery^30^, and a phenotype-driven, whole transcriptome approach to drug discovery can yield additional insights beyond the single receptor- or molecule-level investigation in traditional drug development pipelines. Also, animal models of chronic pain have thus far shown limited utility in nominating new therapeutics^31^. Drug discovery that begins first with human data, in a “bedside to bench” approach, may be additionally informative in the study of chronic pain treatment^10^. Applying this framework of paired genetic and transcriptomic drug repurposing approaches, we nominate FDA-approved medications for further investigation in the treatment of chronic pain.

## Methods

### Signature Mapping

#### Transcriptome-wide association studies

We performed summary-based transcriptome-wide association studies using S-PrediXcan^32^ across 17 blood, skeletal muscle, and central nervous system tissue models built from the Genotype-Tissue Expression Project (GTEx) Project^33^, Depression Genes and Networks (DGN)^34^, the CommonMind Consortium (CMC)^35^, and PsychENCODE^36^ reference transcriptome datasets. The tissue models included: anterior cingulate cortex (BA24), amygdala, caudate (basal ganglia), cerebellar hemisphere, cerebellum, cortex, frontal cortex (BA9), hippocampus, hypothalamus, nucleus accumbens (basal ganglia), putamen (basal ganglia), skeletal muscle, spinal cord (cervical c-1), and substantia nigra GTEx v8 models, CMC dorsolateral prefrontal cortex^37^, DGN whole blood^38^, and PsychENCODE combined temporal and frontal cortex^39^. Tissues were chosen due to previous GWAS enrichment of chronic pain for blood, skeletal muscle, and tissues of the central nervous system through gene-level association testing^3,40,41^. TWAS results were fine-mapped using the FOCUS^42^ tool using tissue-specific reference weight databases imported from each respective PrediXcan model and LD estimated from 1000 Genomes European Phase 3 reference genotypes. We ran FOCUS with all default setting excluding the – p-threshold flag, which was set to a p-value of 0.01 as the minimum GWAS p-value required to perform TWAS fine-mapping.

#### Disease expression signatures

This study included three chronic pain disease expression signatures: 1) presence of chronic pain, 2) severity of chronic pain, and 3) resolution of acute pain. Effective prevention and management of acute pain is an important factor in reducing the likelihood of chronic pain development^43^. Drugs that show similar effects on gene expression as the resolution of acute pain (positive connectivity score) may be additional therapeutic candidates for chronic pain treatment. Presence of chronic pain was defined as the application of S-PrediXcan to summary statistics of a case-control GWAS of chronic multisite pain, with a case defined as an individual with more than one pain site lasting longer than 3 months^4^. Severity of chronic pain was defined as the application of S-PrediXcan to summary statistics of a continuous GWAS of number of chronic pain sites^3^. Expression signatures for resolution of acute pain were taken from a published whole blood differential expression study of acute low back pain over a three month period in patients^21^.

#### Drug expression signatures: Connectivity Map (CMap) Resource

We obtained drug expression signatures from the Connectivity Map (CMap) resource, a compendium of drug compound effects on gene expression across diverse cell lines^22^. The Expanded CMap LINCS Resource 2020 data release was downloaded from https://clue.io/data/CMap2020#LINCS2020 in April 2022. We used level 5 replicate-consensus compound perturbagen signatures, filtering for high quality signatures as defined by the CMap resource as qc_pass = 1 and (median_recall_rank_spearman <=5 or median_recall_rank_wtcs_50 <=5). The final filtered dataset included 89,878 drug signatures across 7,668 compounds and 156 cell types (**Figure S1**).

#### Ensemble Connectivity Score

In the last two decades researchers have proposed many connectivity scores for the comparison of drug and disease gene expression signatures, with few studies comparing the performance of each approach^44–47^. Connectivity scores generally fall into two categories: 1) enrichment tests or 2) similarity measures such as correlation or cosine similarity, and each connectivity score comes with its own limitations. By considering ranks rather than effect estimates, non-parametric enrichment approaches like the modified Kolmogorov-Smirnov test may overemphasize non-differentially expressed genes, leading to an inflated false positive rate, or miss potential associations by not considering the magnitude of differential expression^48^. Conversely, similarity measures are more liable to detection noise due to lowly-expressed genes, and there is no consensus regarding the appropriate number of top and bottom differentially-expressed genes to include in association testing^48^. Further comparison of available connectivity scores can be found in the comprehensive review by Samart et al. (2021)^48^. There is some evidence that combining multiple individual connectivity scores into an ensemble score is effective in nominating drug repurposing candidates and improves performance above individual scores alone^26,47,49^.

In this study we applied an ensemble connectivity score combining five popular individual connectivity scores: the weighted connectivity score^22^ and four eXtreme scores (Xsum, XCor, XSpe, and XCos)^46^. **Figure 1** below provides a flowchart of the connectivity score calculation. TWAS results at four significance thresholds (FDR 5%, 10%, 20% and the 90% fine-mapped credible set) and differential expression study results at three significance thresholds (FDR 1%, 5%, 10%) were included in the connectivity score. The ensemble connectivity score is calculated as follows:

1. Individual connectivity scores (WCS, XSum, XCor, XSpe, XCos) are calculated for each disease signature using Z-scores from disease and drug gene expression signatures.
2. Individual connectivity scores are centered and scaled.
3. Individual scores are averaged across significance thresholds.
4. Mean scores across significance thresholds are averaged across score types to generate the ensemble score.

**Figure 1.**
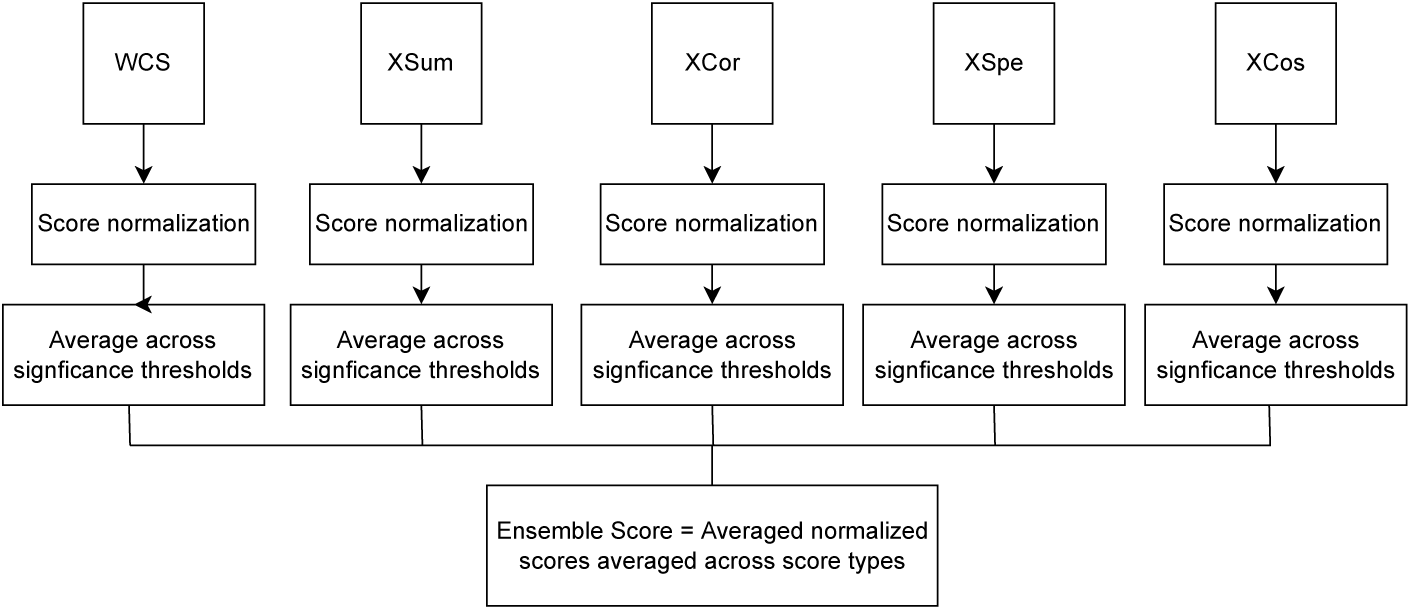
Flowchart outlining calculation of ensemble connectivity score.

We performed permutation testing to understand the range of ensemble scores we would expect due to chance and assign formal significance to our ensemble scores. We scrambled each chronic pain disease signature, randomly reassigning genes to association statistics and again performed the signature mapping analysis across 100 permutations for each tissue. We calculated the empirical p-value as the number of permuted scores less than the test ensemble connectivity score, adding 1 to the numerator and denominator to account for uncertainty in estimation of the p-value^50^.

### Genome-wide association study of chronic pain in Mount Sinai BioMe Biobank

#### GWAS Cohort

We performed genome-wide association studies of chronic pain using the CBIPM-BioMe Biobank Program dataset, hosted under the Mount Sinai Data Ark. This data contains de-identified imputed genotype data and electronic health records. The electronic health records were collected and transformed to the Observational Medical Outcomes Partnership (OMOP) Common Data Model (CDM) format, and includes demographics, encounters, labs, diagnoses, and medications, among other patient information. Further details regarding data acquisition and quality control procedures can be found at the CBIPM-BioMe Program web page.

#### Classification of chronic pain diagnosis

Diagnoses were determined using the Systematic Nomenclature of Medicine-Clinical Terms (SNOMED-CT) terminology, core codes within the OMOP Data Model^51^. A patient required two instances of a particular SNOMED code to be considered to have that condition. We derived two chronic pain phenotypes, one from a list of codes specifically relevant to chronic pain (chronic pain narrow), and a second, broader phenotype including codes of chronic pain and related conditions (chronic pain broad). Chronic pain (narrow) codes include codes characterizing chronic pain or chronic pain syndrome, central pain syndrome, chronic back pain, chronic headache, chronic migraine, complex regional pain syndrome, and phantom limb syndrome with pain. Chronic pain (broad) codes include chronic pain (narrow) conditions as well as diagnoses of Crohn’s disease, ulcerative colitis, arthritis, temporomandibular joint disorders, neuralgia, chronic fatigue syndrome, chronic interstitial cystitis, chronic ulcerative pancolitis, endometriosis, fibromyalgia, irritable bowel syndrome, neuritis, and vulvodynia (**Table S1**).

#### GWAS analysis

SNPs with a minor allele frequency < 0.05, genotyping imputation info score < 0.7, a Hardy-Weinberg equilibrium test p < 10e-8, or missing call rate > 0.05 were removed. Participants with discordant self-reported and genotype-derived sex information, genotype missing rate > 0.05, a heterozygosity rate differing more than 3 SD from the mean, and non-European genotype-derived ancestry were excluded from the analysis. We also identified pairs of related individuals, and excluded one member of each pair with kingship coefficient > 0.0884 using the –king-cutoff parameter of the plink2 software^52^. The final GWAS cohorts included 1,474 individuals diagnosed with narrowly-defined chronic pain, and 5,091 individuals with broadly-defined chronic pain (**Table 1**). Autosomal GWAS for broad and narrowly-defined chronic pain were run using plink –glm function, fitting logistic regression models adjusted for ten genotype PCs, age, sex, and sequencing chip.

**Table 1.**
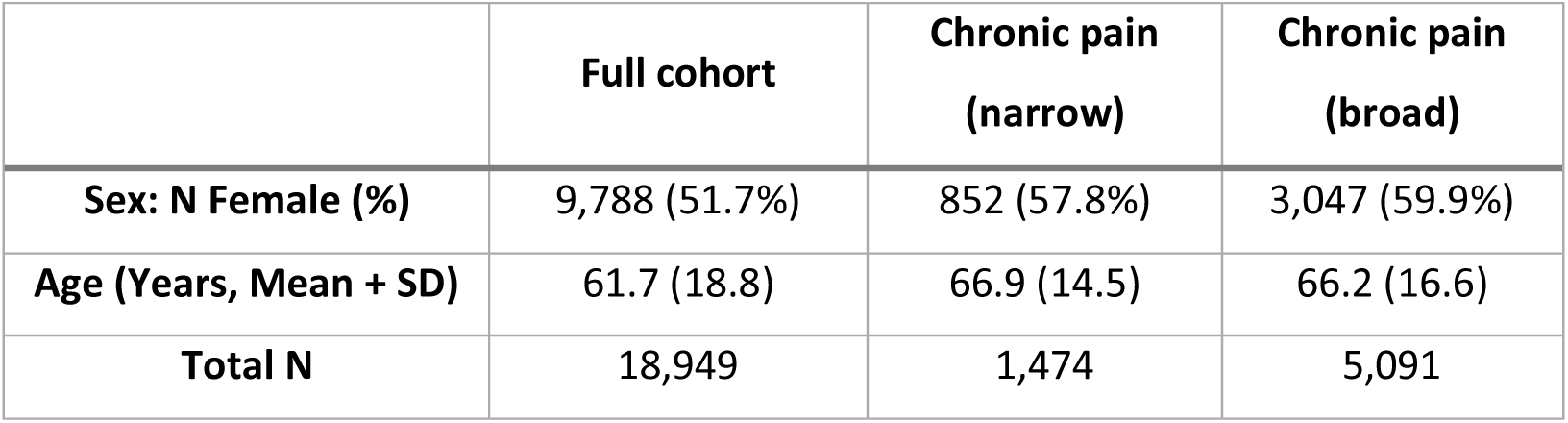
Demographic information for final GWAS cohort in Mount Sinai health system.

#### Genetic correlation analysis

We assessed genetic overlap between the two chronic pain phenotypes used in signature mapping analysis (number of chronic pain sites^3^, presence of chronic multisite pain^4^) and the BioMe narrow chronic pain GWAS phenotype using multivariable LD score regression^53,54^ (LDSC) within the GenomicSEM^55^ R package. Files were munged using info score filter of 0.9 (note info score was not available for all three traits) and a MAF threshold of 0.01. Multivariable LDSC was then performed with sample prevalence parameters 7.7% (BioMe narrow chronic pain GWAS) and 50% (GWAS of presence of chronic multisite pain), and population prevalence as 20% for both (GWAS measuring number of chronic pain sites is quantitative, therefore sample prevalence and population prevalence were set as NA).

### Mendelian randomization analysis

We carried out unidirectional two-sample MR to test for causal effects of using certain medication classes on chronic pain. For this analysis we leveraged 23 previously published medication use GWAS representing 23 medication classes, derived from the UK Biobank cohort, and summary statistics from our own GWAS of ‘narrow’ chronic pain in Mount Sinai BioMe (described in previous section) (**Table 2**).

**Table 2.** GWAS summary statistics used in Mendelian randomization analysis.

| Trait | Source | Cohort | N | N_SNP |
| --- | --- | --- | --- | --- |
| Chronic pain (narrow) | Cote | Mount Sinai BioMe | 18,949 | 7,288,503 |
| Medication use traits | Wu et al<sup>56</sup> | UK Biobank<sup>57,58</sup> | 78,808 -305,913 | 4,653,539 |

N = total sample size, N\_SNP = number of SNPs included in GWAS

We used three MR^59^ methods; MR-Egger^60^, MR-IVW (inverse variance weighted)^61^, and MR-RAPS (robust adjusted profile score)^62^. Both medication use and chronic pain are highly polygenic complex traits, and pleiotropy and other contributors to IV assumption violation are likely. These methods each account for this probably departure from perfect IV assumption adherence (see also Slob & Burgess^63^ for comparative review), increasing power to find potential causal relationships. MR-RAPS also accounts for (and power to find causal effects is increased by) inclusion of ‘weak’ (below genome-wide significance) instruments (SNPs)^62^. To prepare summary statistics for MR, we used the R packages ‘TwoSampleMR’^64,65^, ‘mr.raps’^62^, ‘ieugwasr’^66^ and related 1000 Genomes European LD reference datasets, and ‘genetic.binaRies’^67^ in order to perform LD clumping locally. For each medication use GWAS, we first used the TwoSampleMR function ‘read_exposure_data’ to begin initial processing GWAS summary statistics, followed by reserving SNPs associated with that medication class use at p < 5 × 10^−6^ and then carrying out LD clumping^68^ (clump_kb = 500, clump_r2 = 0.001, clump_p = 0.99) using ‘ieugwasr’ ‘ld_clump’ function, local copies of 1000 Genomes European genetic ancestry plink files, and ‘genetics.binaRies’ ‘get_plink_binary’. We then used ‘TwoSampleMR’ ‘read_outcome_data’ to read in the subset of chronic pain GWAS summary statistics corresponding to the SNPs remaining in the exposure (medication use) summary statistics following clumping. We harmonized using ‘TwoSampleMR’ ‘harmonise_data’ function, and performed MR-Egger and MR-IVW on the final harmonized dataset using ‘TwoSampleMR’ ‘mr’. We then performed MR-RAPS using ‘mr.raps’ ‘mr.raps.all’, retaining any warning messages for examination and then running each of the six models individually with their corresponding functions (see mr.raps vignette for detail) to obtain a p value (since this is not outputted by mr.raps.all). For models with robust loss function (Huber or Tukey) the recommended default k values were used.

MR-RAPS involves fitting of a total of 6 models, and the most appropriate is reserved – in our analyses this was a model without overdispersion (indicating not all instruments are horizontally pleiotropic) and with robust loss function (robust loss functions Huber or Tukey are less sensitive to outliers). This resulted in a total of three MR outputs (Egger, IVW, MR-RAPS robust without overdispersion) per exposure-outcome analysis (23 analyses).

We applied two levels of multiple testing correction: Bonferroni correction within-analysis (threshold 0.05/3 = 0.017) and FDR correction across all analyses (69 analyses total). We consider unadjusted MR p < 0.05 to indicate suggestive significance.

## Results

### Transcriptome-wide association studies reveal pain-related gene expression changes for relevant tissues

There are few well-powered human differential expression studies of chronic pain or pain-related conditions (∼n < 150 cases)^16–20^. Therefore, in order to identify chronic pain-associated genes for downstream signature mapping, we used S-PrediXcan to impute genetically regulated gene expression (GREx) from genotype in two chronic pain phenotype GWAS: presence of chronic multisite pain and number of chronic pain sites^3,4^. We imputed GREx across 17 tissue types with previous chronic pain GWAS enrichment^3,4,40,41^, including tissues of the central nervous system, blood, and skeletal muscle. We identified 2,001 gene-tissue associations (p_fdr_<0.05) with severity of multisite chronic pain across 17 tissues and 1,180 unique genes. We identified 957 gene-tissue associations (p_fdr_<0.05) with presence of chronic multisite pain across 17 tissues and 583 unique genes. We see the largest number of significant gene associations for whole blood, the combined temporal and frontal cortex, skeletal muscle, cerebellum, and the frontal cortex. These results are likely influenced by both the biological relevance of the tissue to chronic pain and the size of the reference panel used to train the transcriptomic imputation model. As expected, we replicate many of the gene-tissue associations identified in a prior summary-based TWAS of chronic pain, including the multi-tissue association of chronic pain with *GMPBB* genetically-regulated expression, and the strong associations of *ECM1* expression and *TARS2* expression with chronic pain in the hippocampus and cerebellum, respectively, among others^69^.

### Transcriptomic signature mapping identifies putative drug candidates for chronic pain

We hypothesized that drugs with an opposite effect on gene expression to chronic pain, or similar effects to the resolution of acute pain, may be potentially therapeutic for the prevention or treatment of chronic pain. To calculate the degree of difference between drug and disease transcriptomic signatures, we applied a novel ensemble score, combining the five most popular single signature mapping scores^22,46^. Our signature mapping analysis nominated 894 putative drug candidates across all three disease signatures (resolution of acute pain: 284, severity of chronic pain: 460, presence of chronic pain: 367)(Presence of chronic pain and number of chronic pain sites: FDR 1%, Resolution of acute pain: FDR 5%) (**Figure 2**; **Table 3**). We chose different significance thresholds to ensure a balanced representation of drug candidates nominated by each disease signature. We observe greater similarity of disease signatures between presence and severity of chronic pain compared to with acute pain resolution (**Figure S2**), and as a result we also see greater overlap in drug candidates between those nominated using the severity or presence of chronic pain TWAS signatures. 25 drug candidates were nominated by all three disease signatures. We see the largest number of significant signature mapping results for transcriptomic signatures of drug perturbagens in A375, PC3, HA1E, and A549 cell lines, as may be expected considering these cell lines contain the greatest proportion of drug signatures in the Connectivity Map resource (**Figure S1**). We also observe significant signature mapping score in more CNS-relevant cell types; for example, neural progenitor cells are among the top five cell lines with the most significant signature mapping results for the resolution of acute pain signature. 210/894 candidates are prescribable in the U.S. (resolution of acute pain: 63, severity of chronic pain: 121, presence of chronic pain: 92). Drug candidates were annotated according to the Anatomical Therapeutic Chemical (ATC) classification system^70^ using the DrugBank Online query tool^71^. Drug candidates span many broad drug classes, including antineoplastic and immunomodulating agents, psychoanaleptics, psycholeptics, drugs related to the genitourinary system and sex hormones, analgesics, and cardiovascular medications, among others (**Table S2**). The overrepresentation of certain drug classes among our signature mapping results is likely affected both by the composition of drug types in the CMap resource and the relevance of the treatment to the biology of chronic pain. Among drugs recommended for chronic pain treatment by the CDC and FDA, and drugs tested in ongoing or completed clinical trials of chronic pain, 13 medications were nominated as drug candidates in our signature mapping analysis: celecoxib, clonazepam, dexamethasone, duloxetine, fluoxetine, lidocaine, nifedipine, omeprazole, paracetamol, pioglitazone, simvastatin, triamcinolone, and varenicline.

**Figure 2.**
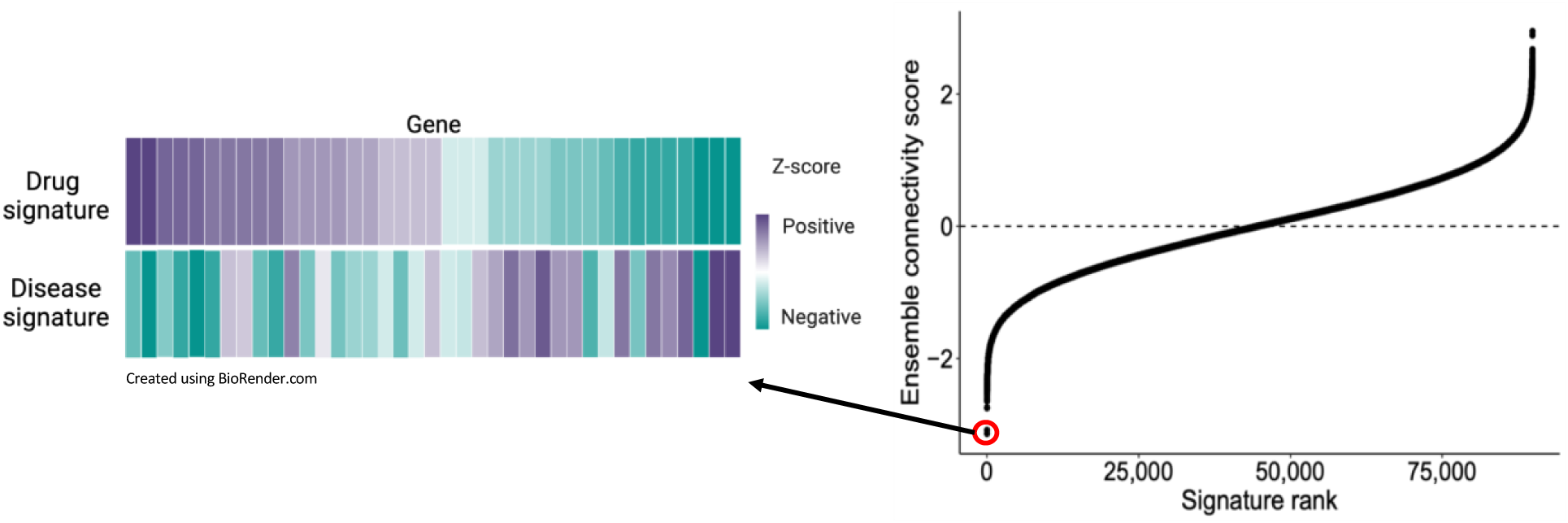
Toy representation of example signature mapping result. Drugs with the opposite effect on gene expression as chronic pain demonstrate negative ensemble connectivity scores. Signature mapping scores typically show a sigmoidal distribution, where a subset of tested medications show strong concordant or opposite effects on gene expression as disease, corresponding to highly positive or negative connectivity scores, respectively.

**Table 3.**
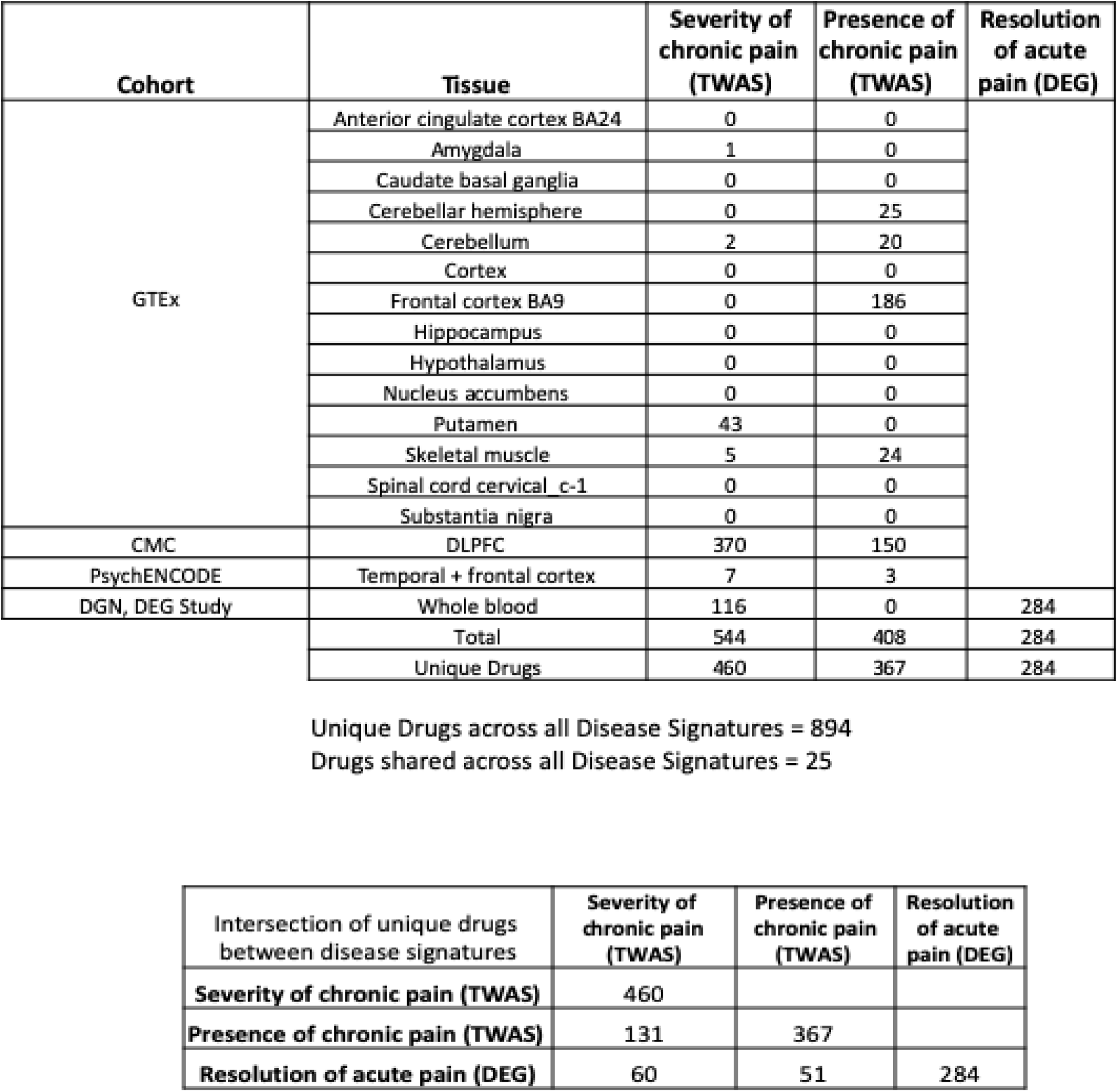
Signature mapping analysis: Number of significant drug candidates by tissue and chronic pain phenotype.

### Causal effects of drug classes on reduced chronic pain in Mendelian randomization analysis

While our signature mapping analysis of chronic pain traits successfully nominated novel drug repurposing candidates in a transcriptomics-informed and tissue-specific context, these drug hypotheses are largely based on associations and do not explicitly imply causal effects of drug exposure on a reduced risk of chronic pain. As a complementary analysis, we used mendelian randomization (MR) to test for potential causal effects of medication use on chronic pain. For this analysis we used previously published medication use GWAS for 23 medication ATC classes in UK Biobank to derive instrumental variables for medication exposure. To derive instrumental variables for the outcome (chronic pain) in a GWAS cohort without overlap with UK Biobank, we performed GWAS of broad (case N= 5,091, control N=13,858) and narrowly-defined (case N= 1,474, control N=17,475) chronic pain in Mount Sinai BioMe.

GWAS for both narrow and broad chronic pain did not reveal any genome-wide significant associations (lowest SNP p-value: narrow = 1.35e-07, broad = 1.96e-06), which is to be expected given the small sample sizes of these cohorts (**Figure S3**). We find greater deviation from the null for the GWAS of narrowly-defined chronic pain, in line with previous research identifying greater power in genetic association studies for more homogeneous disease cohorts^72^, and thus used the “narrow” case definition GWAS for subsequent analysis. Narrow chronic pain, presence of multisite chronic pain^4^, and number of chronic pain sites^3^ all showed significant correlation (**Table S3,S4**).

MR analysis identified three medication classes with a suggestive (p-value<0.05) causal effect on chronic pain (**Table S5**), including N02A (Opioids), B01A (antithrombotic agents), and A02B (drugs for peptic ulcer and GERD)(**Table 4**). N02A is also significantly associated after within-analysis Bonferroni correction (pval < 0.017). Q (heterogeneity) p values all > 0.05, indicating no significant horizontal pleiotropy is present across SNP instruments, and warnings when performing MR-RAPS indicated robust models without overdispersion were most appropriate, meaning SNP instruments tended not to display systematic horizontal pleiotropy but instead idiosyncratic (a small subset of SNPs exhibit horizontal pleiotropy). Results where A02B use is the exposure showed suggestively significant (p < 0.05) negative (beta < 0) effect of this medication on chronic pain (i.e., causing improvement), however the MR Egger intercept was significantly different from 0 (Table S5 p = 0.0053), suggesting significant directional pleiotropy and/or violation of the InSIDE assumption^73^ among SNP instruments – IVW causal estimates are therefore likely biased overall for this drug class and should not be further interpreted. In the case of antithrombotic agents (B01A), the MR Egger intercept is not significantly different from zero (Table S5 p = 0.46), and the MR Egger causal test p value > 0.05, indicating only the IVW and MR-RAPS estimates should be interpreted further. These causal estimates > 0 and are suggestively significant (p > 0.05), suggesting use of this medication has a potentially positive causal effect on chronic pain (i.e., may increase pain or worsen treatment outcomes).

**Table 4:** MR results with suggestive significance.

| Exposure | Method | nSNP | b | se | pval | Q | Q_pval |
| --- | --- | --- | --- | --- | --- | --- | --- |
| **Drugs for peptic ulcer and gastro-esophageal reflux disease (A02B)** | IVW | 34 | -0.237 | 0.182 | 0.193 | 32.80 | 0.48 |
|  | MR Egger |  | -2.311 | 0.938 | 0.019 | 27.72 | 0.68 |
|  | MR-RAPS |  | -0.240 | 0.195 | 0.219 | NA | NA |
| **Antithrombotic agents (B01A)** | IVW | 21 | 0.439 | 0.255 | 0.086 | 28.01 | 0.11 |
|  | MR Egger |  | -0.581 | 0.867 | 0.511 | 25.95 | 0.13 |
|  | MR-RAPS |  | 0.478 | 0.228 | 0.036 | NA | NA |
| **Opioids (N02A)** | IVW | 20 | -0.455 | 0.185 | 0.014 | 23.84 | 0.20 |
|  | MR Egger |  | -0.203 | 1.212 | 0.869 | 23.78 | 0.16 |
|  | MR-RAPS |  | -0.511 | 0.178 | 0.004 | NA | NA |

b = beta value (causal estimate value), se = standard error of beta, pval = p value for beta, Q = heterogeneity measure (heterogeneity of individual SNP causal estimate values), Q\_pval = p value associated with Q. IVW = Inverse variance weighted.

## Discussion

In this study we establish a novel genetics-informed framework to drug repurposing for chronic pain, leveraging existing genetic and transcriptomic data resources. We identify FDA-approved drugs that we hypothesize may be beneficial for prevention or treatment of chronic pain, detected through reversed transcriptomic impacts to chronic pain and association with reduced chronic pain through causal inference approaches. Signature mapping has proven effective in prior studies of other complex conditions such as hyperlipidemia and hypertension^25^, Alzheimer’s disease^24^, and ulcerative colitis^23^, among others. In order to increase predictive power, we extended the current model to include a novel ensemble connectivity score, and signature mapping analyses for multiple pain phenotypes using transcriptomics from both measured and genetically-predicted gene expression. We predict 210 specific drugs, prescribable in the U.S., to have significant reversed effects on gene expression as chronic pain.

Some current pain treatments are supported by our signature mapping analysis, such as duloxetine, an antidepressant prescribed for neuropathic pain relief (resolution of acute pain: score= 2.73, p-value=1.55×10^−5^), and lidocaine, an analgesic that can be administered topically or intravenously for the management of chronic or post-operative pain (Number of chronic pain sites TWAS (whole blood): score= -1.69, p-value=5.26×10^−5^; Presence of chronic multisite pain TWAS (frontal cortex): score= -1.60, p-value= 0.00011)^74^. Among the significant protective drug associations are also various cardiovascular medications such as calcium channel blockers, cardiac glycosides, ACE inhibitors, and beta blockers, as well as corticosteroids, analgesics, and antipsychotic and antidepressant drugs. It is interesting that this genetically-informed drug prioritization approach nominated cardiovascular medications as therapeutic for chronic pain; chronic pain is comorbid with cardiovascular disease, and observational studies provide evidence for a dose-dependent relationship, where greater pain severity is associated with greater cardiovascular disease^75,76^. Family and twin analysis suggests a shared genetic predisposition to chronic pain and cardiovascular disease^77^, and genetic predisposition to multisite chronic pain is associated with a higher risk of cardiovascular disease and related conditions in a recent two-sample Mendelian randomization study, suggesting even a possible causal role of chronic pain in cardiovascular disease development^78^. While further studies are needed to understand the co-occurrence of these conditions and the potential mediating effects of related factors like depression, socioeconomic status, and physical activity, there seems to be a complex interplay between chronic pain and cardiovascular disease, and our analysis supports a possible therapeutic role of cardiovascular medication in chronic pain treatment.

11 medications were both tested in ongoing or completed trials of chronic pain and nominated by our signature mapping analysis. Pioglitazone, an anti-diabetes medication, interestingly has evidence of therapeutic effects in multiple systems, having shown to relieve pain symptoms in both gut- and neuropathic-related mouse pain models^79–81^. We also nominate other medications with interesting preliminary evidence of therapeutic potential for chronic pain treatment. For example the antidepressant mirtazapine has shown preliminary evidence of effectiveness in reducing pain symptoms and pain-related sleep disruption in human and animal studies of chronic pain^82,83^. Atorvastatin and rosuvastatin, cholesterol-lowering medications that act through HMG-CoA reductase inhibition, display anti-inflammatory and pain-reducing effects when tested in rodent models of neuropathic pain^84–87^. Lastly, exposure to atypical antipsychotics such as ziprasidone, quietiapine, clozapine, and olanzapine have been previously investigated as potential secondary chronic pain treatments and all show reversed effects on pain-induced transcriptional changes in our analysis^88,89^. This study provides many genetically-informed future avenues for investigation of alternative pharmaceutical chronic pain treatments.

One advantage of this analytical approach is that signature mapping analysis is largely hypothesis generating and agnostic to prior understanding of disease etiology or current therapeutics, making this approach to drug repurposing particularly useful for the study of diseases of unknown or heterogeneous etiology^90^. Additionally, this study leverages human data for drug candidate testing as opposed to phenotyping exclusively in cell or animal disease models, potentially allowing for more complete modeling of the clinical complexity of chronic pain treatment. Although we may expect not all disease effects to be mediated through changes in gene expression, this framework can be extended as more multi-omic datasets of chronic pain become available and is an informative drug repurposing framework for diseases lacking in effective, long-term drug treatment.

MR analyses highlighted three medication classes with suggestive (p-value< 0.05) causal effects on chronic pain, two of which meet MR assumptions and can be further interpreted. Antithrombotic agents show a potential positive causal effect on chronic pain, meaning exposure to these medications may increase pain or worsen treatment outcomes. Opioids show a negative causal effect on chronic pain, indicating therapeutic benefit, and is the only medication passing within-analysis Bonferroni correction (p-value < 0.017). These findings are broadly in line with previous literature on these drug classes and chronic pain – opioid medications are powerful painkillers ^91,92^, albeit with unclear effectiveness in long term treatment of chronic pain ^15,93^. Antithrombotic agents, including vitamin K antagonists like warfarin, factor Xa inhibitors like rivaroxaban, and heparin, can have pain and chronic pain as side effects through various mechanisms^94^. While this analysis recapitulates some findings from the main drug signature mapping analysis, it also illustrates the utility of signature mapping for specific drug investigation alongside these more overarching and less powerful MR analyses, for finding useful drug therapies in chronic pain.

Results of this study must be interpreted in the context of several limitations. First, the transcriptomic imputation models used to estimate two disease expression signatures in our study are derived from post-mortem tissue, and do not consider environmental, developmental, or long-range genetic effects on gene expression with potential disease relevance. Similarly, while CMap is a comprehensive resource testing many drug compounds, these expression signatures are largely derived from cancer cell lines and likely do not model the complexity of medication effects in vivo, and some drugs may also be tested at dosages or durations that are not clinically feasible. We also note that electronic health record data is not primarily intended for research use. While we made efforts to account for data missingness and incomplete phenotyping by testing specific and broad chronic pain phenotypes, our GWAS of chronic pain and subsequent mendelian randomization findings may still be biased due to issues of incomplete chronic pain diagnosis. Future investigation of chronic pain treatment in the EHR would benefit from more specific follow-up analysis accounting for comorbid conditions or differing therapeutic benefit in chronic pain subtypes. Finally, we would like to emphasize results of this study are based on statistical modeling of population-level genetics, and do not demonstrate any causal effects of drug exposure on protection against development of chronic pain. Extensive follow-up is needed to understand the therapeutic benefit of these medications, including testing of candidate drugs in cell or animal models of chronic pain, as well as evaluation of side effects and safety in long-term treatment.

There are many promising future avenues to further understanding of the effectiveness of these medications in chronic pain treatment. The development of transcriptomic resources in more relevant cellular contexts to chronic pain, as opposed to cancer cell lines, would likely better recapitulate drug treatment effects and yield greater insight into potential repurposing candidates. Also, the chronic pain TWAS used for two of three disease signatures in this study are derived from individuals of European ancestry, and future work would benefit from the inclusion of diverse ancestral groups in the estimation of genetically-informed chronic pain disease expression signatures. Finally, while electronic health record resources from major hospital systems are extensive, longitudinal resources of patient interactions with the healthcare system, datasets with more precise chronic pain patient phenotyping may be additionally informative for accurate diagnosis and causal inference in future genetic studies of chronic pain.

## Data Availability

All data produced in the present study are available upon reasonable request to the authors.

## Acknowledgments

This work was supported in part through the computational and data resources and staff expertise provided by Scientific Computing and Data at the Icahn School of Medicine at Mount Sinai and supported by the Clinical and Translational Science Awards (CTSA) grant UL1TR004419 from the National Center for Advancing Translational Sciences.

