## Supplement for "A computational genetic- and transcriptomics-based study nominates drug repurposing candidates for the treatment of chronic pain"

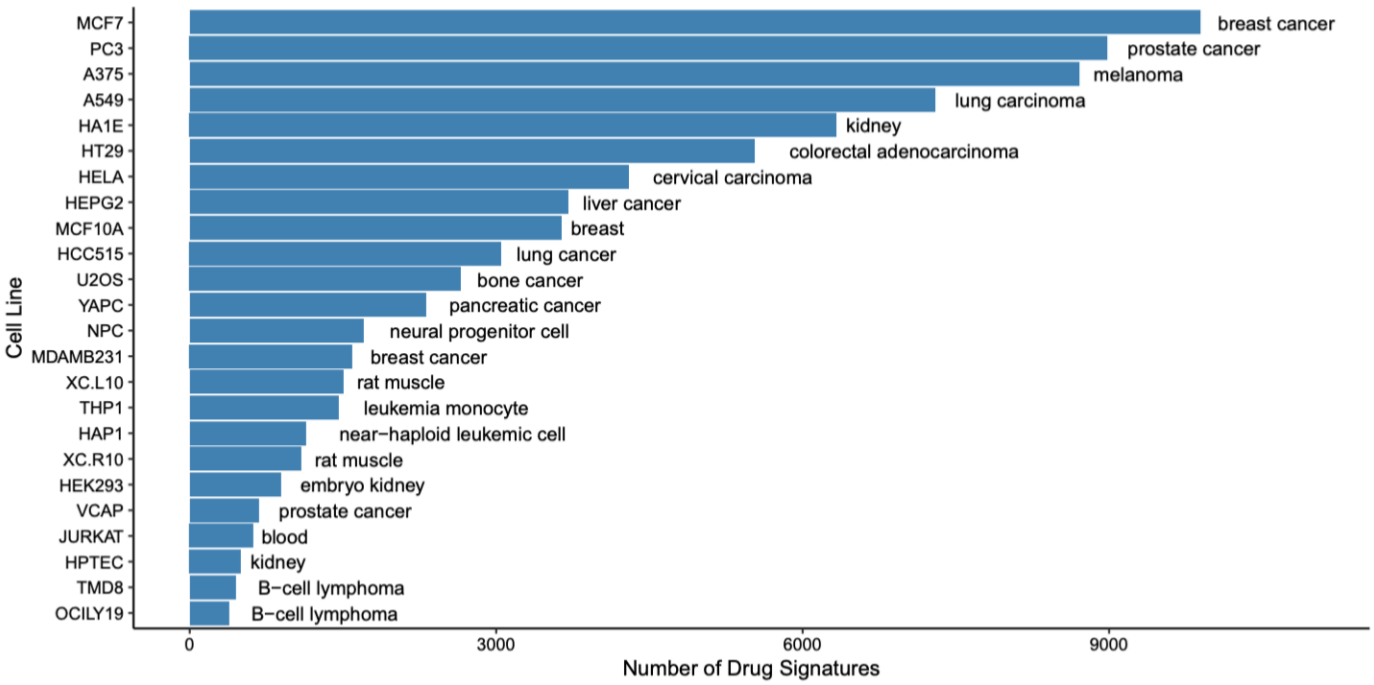


**Figure S1. Number of drug signatures per cell line in the CMap resource, measured across the top 25 cell lines.**

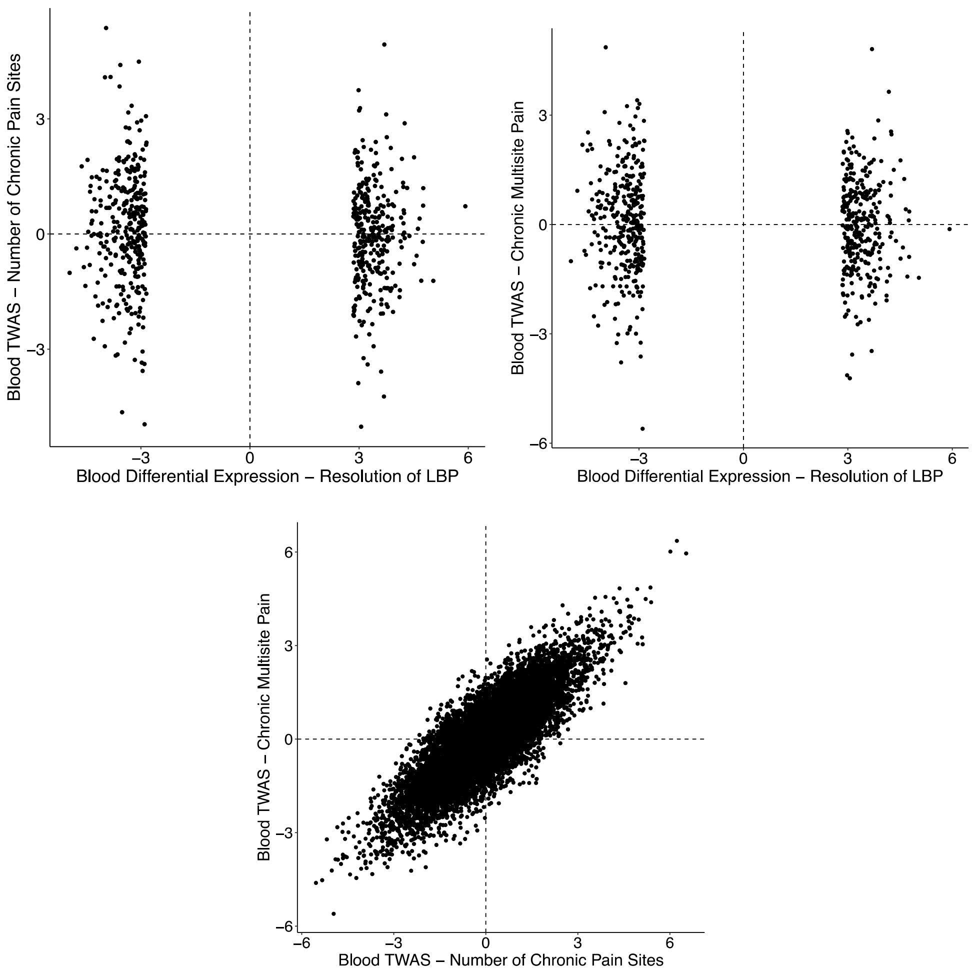


**Figure S2. Comparison of disease expression signatures between chronic pain phenotypes.**

Each point represents a single gene. We see strong similarity between TWAS of the presence of chronic multisite pain and the number of chronic pain sites, while the differential expression signature of acute pain resolution is distinct. LBP = lower back pain.

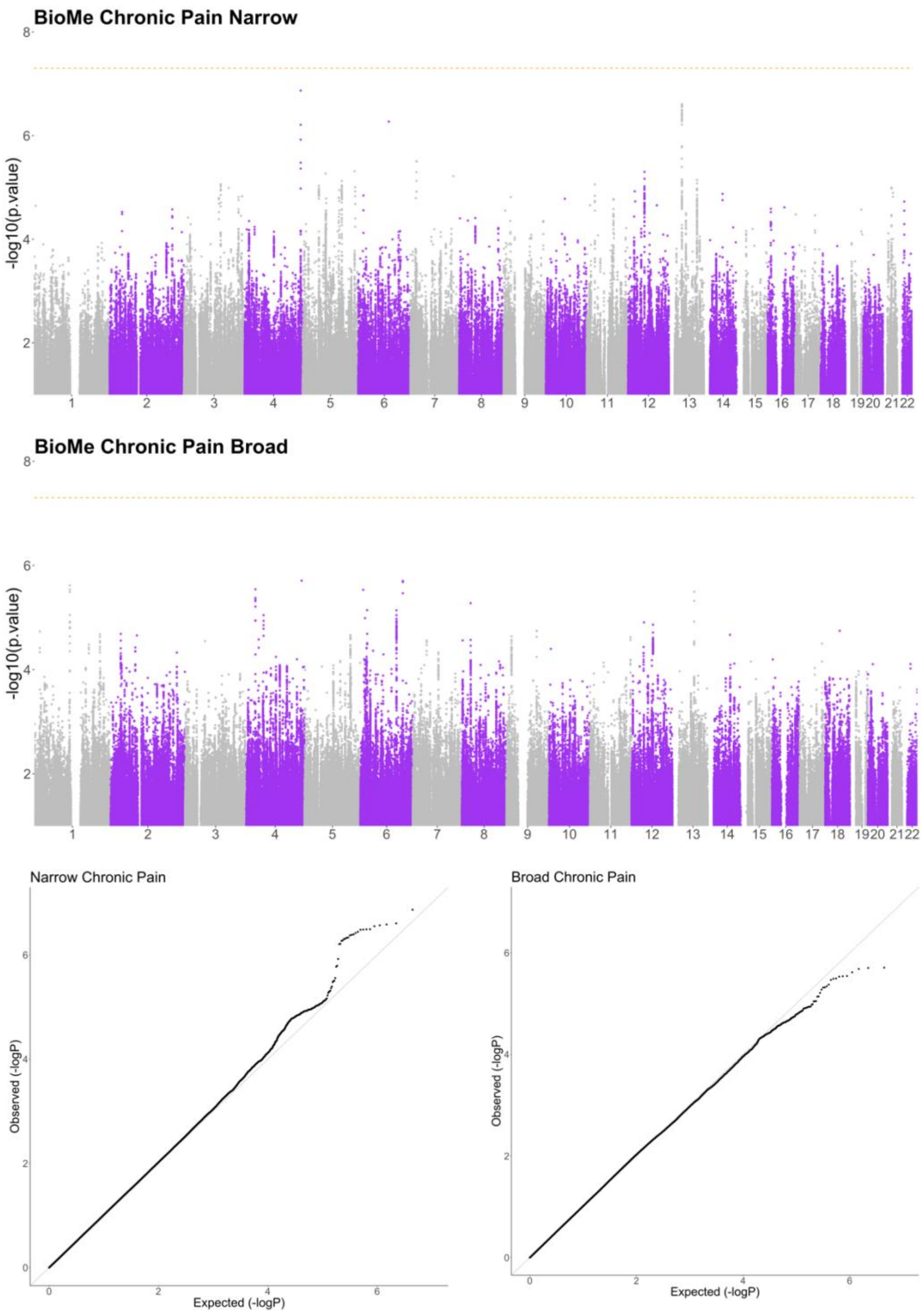


**Figure S3. Manhattan plots and QQ plots for GWAS of narrow and broad chronic pain in the Mount Sinai BioMe dataset.**

**Table S1. Number of patients with each chronic pain condition.**

| SNOMED-CT code | Number of patients |
|---|---|
| Chronic pain* | 1050 |
| Osteoarthritis | 621 |
| Bilateral osteoarthritis of knees | 538 |
| Osteoarthritis of knee | 527 |
| Osteoarthritis of hip | 440 |
| Arthritis | 402 |
| Osteoarthritis of right knee joint | 400 |
| Chronic fatigue syndrome | 391 |
| Irritable bowel syndrome | 369 |
| Osteoarthritis of left knee joint | 354 |
| Chronic low back pain* | 346 |
| Low back pain co-occurrent with neuralgia of left sciatic nerve | 300 |
| Ulcerative colitis | 292 |
| Crohn's disease | 287 |
| Idiopathic osteoarthritis | 267 |
| Localized primary osteoarthritis of the shoulder region | 254 |
| Chronic pain syndrome* | 228 |
| Fibromyalgia | 223 |
| Complication due to ulcerative colitis | 209 |
| Rheumatoid arthritis | 201 |
| Complication due to Crohn's disease | 179 |
| Crohn's disease of small intestine | 162 |
| Localized primary osteoarthritis of the hand | 161 |
| Crohn's disease of small AND large intestines | 158 |
| Irritable bowel syndrome characterized by constipation | 156 |
| Chronic ulcerative pancolitis | 142 |
| Localized primary osteoarthritis of the ankle and/or foot | 140 |
| Osteoarthritis of right hip joint | 137 |
| Irritable bowel syndrome with diarrhea | 133 |
| Complication due to Crohn's disease of small and large intestines | 132 |
| Psoriatic arthritis | 123 |
| Crohn's disease of large bowel | 116 |
| Ulcerative pancolitis | 116 |
| Complication due to Crohn's disease of small intestine | 113 |
| Osteoarthritis of left hip joint | 106 |
| Complication due to chronic ulcerative pancolitis | 103 |

| | |
|---|---|
| Thoracic and lumbosacral neuritis | 98 |
| Complication due to Crohn's disease of large intestine | 93 |
| Chronic migraine without aura with status migrainosus* | 90 |
| Seropositive rheumatoid arthritis | 81 |
| Fistula of intestine due to Crohn's disease of small and large intestine | 78 |
| Chronic tension-type headache* | 72 |
| Rectal hemorrhage due to chronic ulcerative pancolitis | 72 |
| Cervico-occipital neuralgia | 67 |
| Chronic migraine without aura* | 67 |
| Endometriosis (clinical) | 65 |
| Rectal hemorrhage due to ulcerative colitis | 63 |
| Trigeminal neuralgia | 62 |
| Chronic intractable migraine without aura* | 56 |
| Seronegative rheumatoid arthritis | 55 |
| Intestinal obstruction due to Crohn's disease of small intestine | 52 |
| Optic neuritis | 52 |
| Postherpetic neuralgia | 50 |
| Rheumatoid factor positive rheumatoid arthritis | 50 |
| Temporomandibular joint disorder | 49 |
| Localized primary osteoarthritis | 48 |
| Irritable bowel syndrome characterized by alternating bowel habit | 45 |
| Fistula of small intestine due to Crohn's disease | 40 |
| Hyperuricemia without signs of inflammatory arthritis and tophaceous disease | 39 |
| Arthritis of spine | 37 |
| Post-herpetic neuritis | 37 |
| Arthritis of shoulder region joint | 33 |
| Fistula of large intestine due to Crohn's disease | 33 |
| Intestinal obstruction due to Crohn's disease of small and large intestine | 33 |
| Chronic interstitial cystitis | 32 |
| Rheumatoid arthritis of multiple joints | 27 |
| Intestinal obstruction due to Crohn's disease | 25 |
| Arthritis of knee | 22 |
| Chronic back pain* | 22 |
| Localized primary osteoarthritis of the wrist | 22 |
| Rheumatoid arthritis of shoulder | 21 |
| Left sided ulcerative colitis | <20 |
| Post traumatic osteoarthritis | <20 |
| Rectal hemorrhage due to Crohn's disease of large intestine | <20 |
| Rectal hemorrhage due to Crohn's disease of small and large intestines | <20 |
| Chronic cluster headache* | <20 |

| | |
|---|---|
| Complex regional pain syndrome of lower limb* | <20 |
| Endometriosis of uterus | <20 |
| Enteropathic arthritis | <20 |
| Suppurative arthritis | <20 |
| Abscess of intestine co-occurrent and due to Crohn's disease of small and large intestine | <20 |
| Localized secondary osteoarthritis of the shoulder region | <20 |
| Rectal hemorrhage due to Crohn's disease | <20 |
| Vulvodynia | <20 |
| Abscess of intestine co-occurrent and due to Crohn's disease of small intestine | <20 |
| Chronic pain due to injury* | <20 |
| Complex regional pain syndrome of upper limb* | <20 |
| Reactive arthritis triad | <20 |
| Rectal hemorrhage due to Crohn's disease of small intestine | <20 |
| Localized primary osteoarthritis of elbow | <20 |
| Polyneuritis | <20 |
| Intestinal obstruction due to Crohn's disease of large intestine | <20 |
| Mononeuritis multiplex | <20 |
| Abscess of intestine co-occurrent and due to Crohn's disease | <20 |
| Abscess of intestine co-occurrent and due to Crohn's disease of large intestine | <20 |
| Juvenile rheumatoid arthritis | <20 |
| Lyme arthritis | <20 |
| Monoarthritis of knee | <20 |
| Arthritis of temporomandibular joint | <20 |
| Central pain syndrome* | <20 |
| Chronic post-traumatic headache* | <20 |
| Complex regional pain syndrome type II upper limb* | <20 |
| Complex regional pain syndrome type II lower limb* | <20 |
| Complex regional pain syndrome* | <20 |
| Fistula of intestine due to ulcerative colitis | <20 |
| Phantom limb syndrome with pain* | <20 |
| Rheumatoid arthritis of wrist | <20 |
| Staphylococcal arthritis | <20 |
| Systemic onset juvenile chronic arthritis | <20 |
| Bacterial arthritis | <20 |
| Fistula of intestine due to chronic ulcerative pancolitis | <20 |
| Intestinal obstruction due to chronic ulcerative pancolitis | <20 |
| Intestinal obstruction due to ulcerative colitis | <20 |
| Monoarthritis | <20 |
| Neuralgia | <20 |

| | |
|---|---|
| Post-herpetic trigeminal neuralgia | <20 |
| Psoriatic arthritis mutilans | <20 |
| Arthritis of hand | <20 |
| Arthritis of left knee caused by bacteria | <20 |
| Arthritis of left temporomandibular joint | <20 |
| Bacterial arthritis of hip | <20 |
| Endometriosis of ovary | <20 |
| Gouty arthritis of the ankle and/or foot | <20 |
| Gouty arthritis of the shoulder region | <20 |
| Knee pyogenic arthritis | <20 |
| Monoarthritis of hand | <20 |
| Monoarthritis of wrist | <20 |
| Osteoarthritis of hip due to dysplasia | <20 |
| Pauciarticular juvenile rheumatoid arthritis | <20 |
| Rheumatoid arthritis - ankle and/or foot | <20 |
| Rheumatoid arthritis - hand joint | <20 |
| Rheumatoid arthritis of hip | <20 |

*Chronic pain (narrow) code, all other codes included in chronic pain (broad) definition.

**Table S2. Tier 1 and 2 ATC classification of drug candidates**

| ATC Level 1 | ATC Level 2 | Count |
|---|---|---|
| alimentary tract and metabolism (7) | Anabolic steroids for systemic use | 1 |
| | Antiemetics and antinauseants | 1 |
| | Drugs used in diabetes | 3 |
| | other alimentary tract and metabolism products | 1 |
| antiinfectives for systemic use (7) | antibacterials for systemic use | 4 |
| | antimycobacterials | 1 |
| | antivirals for systemic use | 4 |
| antineoplastic and immunomodulating agents (74) | antineoplastic agents | 63 |
| | endocrine therapy | 4 |
| | immunosuppressants | 4 |
| | other | 3 |
| antiparasitic products, insecticides and repellants (2) | anthelmintics | 1 |
| | antiprotozoals | 1 |
| blood and blood forming organs (5) | antihemorrhagics | 1 |
| | antithrombotic agents | 4 |
| cardiovascular system (19) | agents acting on the renin-angiotensin system | 5 |
| | antihypertensives | 3 |
| | beta blocking agents | 2 |
| | calcium channel blockers | 1 |
| | cardiac therapy | 2 |
| | diuretics | 3 |
| | lipid modifying agents | 1 |
| | peripheral vasodilators | 1 |
| | vasoprotectives | 1 |
| dermatologicals (6) | antibiotics and chemotherapeutics for dermatological use | 3 |
| | antifungals for dermatological use | 2 |
| | corticosteroids, dermatological preparations | 1 |
| genito urinary system and sex hormones (11) | sex hormones and modulators of the genital system | 8 |
| | urologicals | 3 |
| musculo-skeletal system (2) | antigout preparations | 1 |
| | other | 1 |
| nervous system (33) | analgesics | 4 |
| | Anti-Parkinson drugs | 1 |
| | antiepileptics | 3 |
| | other nervous system drugs | 2 |
| | psychoanaleptics | 12 |
| | psycholeptics | 11 |
| respiratory system (3) | antihistamines for systemic use | 1 |
| | cough and cold preparations | 1 |

| | | |
|---|---|---|
| | other | 1 |
| sensory organs (1) | ophthalmologicals | 1 |
| systemic hormonal preparations, excl. sex hormones and insulin (3) | calcium homeostasis | 1 |
| | pituitary and hypothalamic hormones and analogues | 1 |
| | corticosteroids for systemic use | 1 |
| various (1) | all other therapeutic products | 1 |
| Unclassified or multiple classifications (35) | | |

**Table S3: Genetic correlation between pain traits**. Standard error of genetic correlation estimate is given in parentheses.

| | Chronic pain (narrow) | Presence of chronic multisite pain | Number of chronic pain sites |
|---|---|---|---|
| Chronic pain (narrow) | 1 | 1.009 (0.306) | 0.97 (0.235) |
| Presence of chronic multisite pain | | 1 | 0.98 (0.0399) |
| Number of chronic pain sites | | | 1 |

**Table S4: Additional trait information:** SNP-h2 = SNP heritability. (SE) = standard error of SNP-h2.

| **Trait** | **Liability scale SNP-h2 (SE)** | **N GWAS** | **Sample prevalence** |
|---|---|---|---|
| Chronic pain (narrow) | 0.044 (0.13) | 18,949 | 0.077 |
| Presence of chronic multisite pain | 0.17 (0.0081) | 164,778 | 0.5 |
| Number of chronic pain sites | 0.0741 (0.0028) | 387,649 | NA |

**Table S5: Mendelian randomization analysis of medication use effects on chronic pain.**

| Exposure | Method | nSNP | **b** | **se** | **pval** | **Q** | **Q_pval** | **pval.fdr** | **Intercept p** |
|---|---|---|---|---|---|---|---|---|---|
| **Drugs for peptic ulcer and gastro-esophageal reflux disease (A02B)** | IVW | 34 | -0.237 | 0.182 | 0.193 | 32.80 | 0.48 | 0.884 | NA |
| | MR Egger | | -2.311 | 0.938 | 0.019 | 27.72 | 0.68 | 0.443 | 0.005 |
| | MR-RAPS | | -0.240 | 0.195 | 0.219 | NA | NA | 0.884 | NA |
| **Drugs used in diabetes (A10)** | IVW | 94 | -0.008 | 0.053 | 0.874 | 96.39 | 0.38 | 0.933 | NA |
| | MR Egger | | -0.050 | 0.126 | 0.695 | 96.25 | 0.36 | 0.933 | 0.719 |
| | MR-RAPS | | -0.009 | 0.054 | 0.867 | NA | NA | 0.933 | NA |
| **Antithrombotic agents (B01A)** | IVW | 21 | 0.439 | 0.255 | 0.086 | 28.01 | 0.11 | 0.845 | NA |
| | MR Egger | | -0.581 | 0.867 | 0.511 | 25.95 | 0.13 | 0.922 | 0.463 |
| | MR-RAPS | | 0.478 | 0.228 | 0.036 | NA | NA | 0.613 | NA |
| **Vasodilators used in cardiac diseases (C01D)** | IVW | 14 | 0.128 | 0.104 | 0.219 | 12.98 | 0.45 | 0.884 | NA |
| | MR Egger | | -0.015 | 0.344 | 0.966 | 12.77 | 0.39 | 0.998 | 0.46 |
| | MR-RAPS | | 0.146 | 0.112 | 0.193 | NA | NA | 0.884 | NA |
| **Antihypertensives (C02)** | IVW | 10 | -0.127 | 0.143 | 0.375 | 10.08 | 0.34 | 0.884 | NA |
| | MR Egger | | 0.283 | 0.649 | 0.674 | 9.58 | 0.30 | 0.933 | 0.535 |
| | MR-RAPS | | -0.157 | 0.145 | 0.279 | NA | NA | 0.884 | NA |
| **Diuretics (C03)** | IVW | 158 | 0.069 | 0.066 | 0.298 | 149.17 | 0.66 | 0.884 | NA |
| | MR Egger | | 0.434 | 0.229 | 0.060 | 146.39 | 0.70 | 0.825 | 0.056 |
| | MR-RAPS | | 0.053 | 0.070 | 0.445 | NA | NA | 0.903 | NA |
| **Beta blocking agents (C07)** | IVW | 102 | 0.094 | 0.084 | 0.264 | 90.91 | 0.75 | 0.884 | NA |
| | MR Egger | | -0.408 | 0.316 | 0.199 | 88.20 | 0.79 | 0.884 | 0.105 |
| | MR-RAPS | | 0.078 | 0.089 | 0.382 | NA | NA | 0.884 | NA |
| **Calcium channel blockers (C08)** | IVW | 161 | 0.057 | 0.065 | 0.382 | 154.38 | 0.61 | 0.884 | NA |
| | MR Egger | | 0.184 | 0.223 | 0.410 | 154.02 | 0.60 | 0.884 | 0.389 |
| | MR-RAPS | | 0.049 | 0.069 | 0.472 | NA | NA | 0.922 | NA |
| | IVW | 271 | 0.115 | 0.065 | 0.077 | 272.88 | 0.44 | 0.845 | NA |
| | MR Egger | | 0.121 | 0.208 | 0.561 | 272.88 | 0.42 | 0.922 | 0.953 |

| | | | | | | | | | |
|---|---|---|---|---|---|---|---|---|---|
| **Agents acting on the renin-angiotensin system (C09)** | MR-RAPS | | 0.110 | 0.068 | 0.107 | NA | NA | 0.884 | NA |
| **HMG CoA reductase inhibitors (C10AA)** | IVW | 131 | -0.039 | 0.093 | 0.678 | 147.00 | 0.15 | 0.933 | NA |
| | MR Egger | | -0.084 | 0.218 | 0.701 | 146.94 | 0.13 | 0.933 | 0.817 |
| | MR-RAPS | | -0.086 | 0.091 | 0.344 | NA | NA | 0.884 | NA |
| **Thyroid preparations (H03A)** | IVW | 189 | -0.011 | 0.039 | 0.783 | 181.75 | 0.61 | 0.933 | NA |
| | MR Egger | | 0.028 | 0.080 | 0.725 | 181.44 | 0.60 | 0.933 | 0.735 |
| | MR-RAPS | | -0.010 | 0.041 | 0.807 | NA | NA | 0.933 | NA |
| **Immunosuppressants (L04)** | IVW | 13 | 0.067 | 0.079 | 0.399 | 5.43 | 0.94 | 0.884 | NA |
| | MR Egger | | 0.078 | 0.260 | 0.771 | 5.43 | 0.91 | 0.933 | 0.951 |
| | MR-RAPS | | 0.066 | 0.084 | 0.430 | NA | NA | 0.899 | NA |
| **Anti-inflammatory and antirheumatic products, non-steroids (M01A)** | IVW | 38 | 0.005 | 0.205 | 0.979 | 47.51 | 0.12 | 0.998 | NA |
| | MR Egger | | 0.216 | 1.104 | 0.846 | 47.46 | 0.10 | 0.933 | 0.847 |
| | MR-RAPS | | -0.048 | 0.192 | 0.803 | NA | NA | 0.933 | NA |
| **Drugs affecting bone structure and mineralization (M05B)** | IVW | 24 | -0.015 | 0.100 | 0.879 | 24.86 | 0.36 | 0.933 | NA |
| | MR Egger | | -0.001 | 0.502 | 0.998 | 24.86 | 0.30 | 0.998 | 0.977 |
| | MR-RAPS | | -0.001 | 0.103 | 0.992 | NA | NA | 0.998 | NA |
| **Opioids (N02A)** | IVW | 20 | -0.455 | 0.185 | 0.014 | 23.84 | 0.20 | 0.443 | NA |
| | MR Egger | | -0.203 | 1.212 | 0.869 | 23.78 | 0.16 | 0.933 | 0.836 |
| | MR-RAPS | | -0.511 | 0.178 | 0.004 | NA | NA | 0.276 | NA |
| **Salicylic acid and derivatives (N02BA)** | IVW | 20 | 0.124 | 0.213 | 0.559 | 20.21 | 0.38 | 0.922 | NA |
| | MR Egger | | 0.425 | 1.101 | 0.704 | 20.13 | 0.33 | 0.933 | 0.826 |
| | MR-RAPS | | 0.133 | 0.220 | 0.545 | NA | NA | 0.922 | NA |
| **Anilides (N02BE)** | IVW | 31 | -0.133 | 0.204 | 0.515 | 27.55 | 0.59 | 0.922 | NA |
| | MR Egger | | -0.233 | 0.861 | 0.789 | 27.53 | 0.54 | 0.933 | 0.9 |
| | MR-RAPS | | -0.132 | 0.217 | 0.544 | NA | NA | 0.922 | NA |

| | | | | | | | | | |
|---|---|---|---|---|---|---|---|---|---|
| **Antimigraine preparations (N02C)** | IVW | 23 | 0.032 | 0.070 | 0.649 | 18.89 | 0.65 | 0.933 | NA |
| | MR Egger | | -0.068 | 0.348 | 0.846 | 18.80 | 0.60 | 0.933 | 0.779 |
| | MR-RAPS | | 0.036 | 0.074 | 0.631 | NA | NA | 0.933 | NA |
| **Antidepressants (N06A)** | IVW | 22 | -0.209 | 0.230 | 0.362 | 23.43 | 0.32 | 0.884 | NA |
| | MR Egger | | -0.830 | 0.919 | 0.377 | 22.87 | 0.30 | 0.884 | 0.443 |
| | MR-RAPS | | -0.264 | 0.234 | 0.259 | NA | NA | 0.884 | NA |
| **Adrenergics, inhalants (R03A)** | IVW | 78 | -0.084 | 0.078 | 0.286 | 88.24 | 0.18 | 0.884 | NA |
| | MR Egger | | -0.175 | 0.208 | 0.403 | 87.98 | 0.16 | 0.884 | 0.621 |
| | MR-RAPS | | -0.084 | 0.077 | 0.272 | NA | NA | 0.884 | NA |
| **Glucocorticoids (R03BA)** | IVW | 41 | -0.035 | 0.090 | 0.699 | 43.30 | 0.33 | 0.933 | NA |
| | MR Egger | | 0.086 | 0.290 | 0.768 | 43.09 | 0.30 | 0.933 | 0.662 |
| | MR-RAPS | | -0.048 | 0.091 | 0.599 | NA | NA | 0.933 | NA |
| **Antihistamines for systemic use (R06A)** | IVW | 20 | 0.090 | 0.130 | 0.490 | 14.07 | 0.78 | 0.922 | NA |
| | MR Egger | | 0.550 | 0.551 | 0.332 | 13.33 | 0.77 | 0.884 | 0.331 |
| | MR-RAPS | | 0.118 | 0.139 | 0.397 | NA | NA | 0.884 | NA |
| **Antiglaucoma preparations and miotics (S01E)** | IVW | 33 | 0.076 | 0.062 | 0.214 | 28.39 | 0.65 | 0.884 | NA |
| | MR Egger | | -0.082 | 0.315 | 0.795 | 28.13 | 0.61 | 0.933 | 0.593 |
| | MR-RAPS | | 0.064 | 0.065 | 0.328 | NA | NA | 0.884 | NA |

b = beta value (causal estimate value), se = standard error of beta, pval = p value for beta, Q = heterogeneity measure (heterogeneity of individual SNP causal estimate values), Q_pval = p value associated with Q. IVW = Inverse variance weighted. Intercept p = p value for MR Egger intercept test (is the intercept significantly different from zero).
